# Attitudes towards booster, testing and isolation, and their impact on COVID-19 response in winter 2022/2023 in France, Belgium, and Italy

**DOI:** 10.1101/2022.12.30.22283726

**Authors:** Giulia de Meijere, Eugenio Valdano, Claudio Castellano, Marion Debin, Charly Kengne-Kuetche, Clément Turbelin, Harold Noël, Joshua Weitz, Daniela Paolotti, Lisa Hermans, Niel Hens, Vittoria Colizza

## Abstract

European countries are focusing on testing, isolation, and boosting strategies to counter the 2022/2023 winter surge due to Omicron subvariants. However, widespread pandemic fatigue and limited compliance potentially undermine mitigation efforts. To establish a baseline for interventions, we ran a multicountry survey to assess respondents’ willingness to receive booster vaccination and comply with testing and isolation mandates. The vast majority of survey participants (N=4,594) was willing to adhere to testing (>91%) and rapid isolation (>88%) across the three countries. Pronounced differences emerged in the declared senior adherence to booster vaccination (73% in France, 94% in Belgium, 86% in Italy). Next, we inferred the vaccine-induced population immunity profile at the winter start from prior vaccination data, immunity waning, and declared booster uptake. Integrating survey and estimated immunity data in a branching process epidemic spreading model, we evaluated the effectiveness and costs of current protocols in France, Belgium, and Italy to manage the winter wave. Model results estimate that testing and isolation protocols would confer significant benefit in reducing transmission (17-24%) with declared adherence. Achieving a mitigating level similar to the French protocol, the Belgian protocol would require 30% fewer tests and avoid the long isolation periods of the Italian protocol (average of 6 days vs. 11). A cost barrier to test would significantly decrease adherence in France and Belgium, undermining protocols’ effectiveness. Simpler mandates for isolation may increase awareness and actual compliance, reducing testing costs, without compromising mitigation. High booster vaccination uptake remains key for the control of the winter wave.

## INTRODUCTION

As the COVID-19 pandemic approaches its third winter season in Europe, the World Health Organization recommends that countries should strategically prepare for a possible surge in cases, hospitalizations, and deaths^1^. Given the largely immunized population, the response needs to adapt to a shifting phase of the pandemic^1–4^.

Contact tracing and quarantine have been progressively eased due to their costs and disruption on the workforce and individual life^5,6^. Case isolation, however, remains key to reduce SARS-CoV-2 spread and protect the health system. Countries adopted different lengths of isolation, with some cutting it short if symptoms subside or the person tests negative (**Table 1**). Vaccination with an additional booster dose is currently ongoing for the fall 2022 campaign^7^. But it is hard to anticipate its uptake, after several vaccine doses and the marked decline in coverage reported for the booster doses administered in the summer 2022 to vulnerable individuals (median uptake across European countries of 11.6% among 60+ by early July 2022^7^).

**Table 1.** Protocols for testing, tracing and isolation adopted in France, Belgium, and Italy in 2022.

|  | France | Belgium | Italy |
| --- | --- | --- | --- |
| *Winter 2021/2022 protocols* | **January-February 2022**<br>**If symptomatic:** vaccinated and unvaccinated individuals test and isolate, if positive.<br>**If traced:** vaccinated individuals test on days 0, 2 and 4 from tracing and isolate if positive; unvaccinated individuals quarantine for 7 days with exit test (if positive, exit on day 10).<br>**Isolation\*:** vaccinated individuals isolate for 5 days with exit test (if positive, exit on day 7); unvaccinated individuals quarantine for 7 days with exit test (if positive, exit on day 10). | **January - April 2022**<br>**If symptomatic:** vaccinated and unvaccinated individuals test and isolate, if positive.<br>**If traced:** vaccinated individuals do not need to quarantine nor to test; unvaccinated individuals quarantine for 10 days, with daily exit tests starting from day 7 (partially vaccinated individuals quarantine for 7 days, with daily exit tests starting from day 4).<br>**Isolation:** vaccinated and unvaccinated individuals isolate for 7 days. | **December 2021 - March 2022**<br>**If symptomatic:** vaccinated and unvaccinated individuals test and isolate, if positive.<br>**If traced:** vaccinated individuals test only if they develop symptoms (test on day 0 and on day 5 from symptom onset) and isolate, if positive; unvaccinated individuals quarantine for 10 days with exit test (if positive, test again until negative).<br>**Isolation:** vaccinated individuals isolate for 7 days with exit test (if positive, test again on day 14, if positive, exit on day 21); unvaccinated individuals quarantine for 10 days with exit test (if positive, exit on day 21). |
| *Spring 2022 protocols* | **Since March 2022 (still in place)**<br>**If symptomatic:** vaccinated and unvaccinated individuals test and isolate, if positive.<br>**If traced:** vaccinated and unvaccinated individuals test on day 2 and isolate, if positive.<br>**Isolation\*:** vaccinated individuals isolate for 5 days with exit test (if positive, exit on day 7); unvaccinated individuals isolate for 7 days with exit test (if positive, exit on day 10). | **Since May 2022 (still in place)**<br>**If symptomatic:** vaccinated and unvaccinated individuals test and isolate, if positive.<br>**If traced:** vaccinated and unvaccinated individuals do not test nor quarantine .<br>**Isolation:** vaccinated and unvaccinated individuals isolate for 7 days. | **Since April 2022 (new one since September 2022\*\*)**<br>**If symptomatic:** vaccinated and unvaccinated individuals test and isolate, if positive.<br>**If traced:** vaccinated and unvaccinated individuals test only if they develop symptoms (test on day 0 and day 5 from symptom onset) and isolate, if positive.<br>**Isolation:** vaccinated individuals isolate for 7 days with exit test (if positive, test again on day 14, if positive, exit on day 21); unvaccinated individuals isolate for 10 days with exit test (if positive, exit on day 21).<br>**\*\* Isolation:** vaccinated and unvaccinated individuals isolate for 5 days with exit test (if positive, test again until negative or exit on day 14) |

\*The requirement on isolation exit based on the presence or absence of symptoms is not mentioned in the table as it cannot be accounted for by the model.

In this study we evaluated the expected performance of the testing and isolation strategies currently adopted in France, Belgium, and Italy to manage COVID-19 through winter 2022/2023. We inferred the vaccine-induced immunity profile of the population of the three countries at the start of the winter season based on vaccination data and accounting for immunity waning. We ran a multi-country survey to anticipate individual behavior from 4,594 survey participants from France, Belgium, and Italy on booster vaccination, adherence to testing and isolation, under a range of protocols (isolation of different durations, with or without exit tests) and conditions (in the presence or absence of recommendations, with free access to tests or at a given cost). We then used a mathematical model of SARS-CoV-2 infection dynamics parameterized to Omicron subvariants and to survey results on expected individual compliance to evaluate the performance of the testing and isolation protocols adopted by the three countries.

## RESULTS

### Attitudes towards vaccination, testing, and isolation in the 2022/2023 winter season

We used the three national platforms of the digital surveillance system Influenzanet^8–10^ in France, Belgium, and Italy (GrippeNet.fr/COVIDnet.fr, Infectieradar.be, Influweb.it, respectively) to investigate prospective behavior on booster uptake, testing, and isolation in the 2022/2023 winter season. Data collection took place in the summer 2022, resulting in 3,534 respondents in France, 741 in Belgium, and 319 in Italy, after accounting for the inclusion criteria (see Methods). In the following, results for the three countries will be listed according to decreasing sample size. After adjusting for age, gender, and education, countries displayed a marked difference in the participants’ propensity to receive an additional booster dose in the fall of 2022 (72.5% of the elderly in France, vs. 93.9% and 86.4% in Belgium and Italy, respectively; **Figure 1A**). Prospective testing behavior showed very high adherence to testing if symptomatic (92.7% in France, 91.2% in Belgium, 97.3% in Italy), and 8 to 15 percentage points lower adherence if asymptomatic (78.2% in France, 82.3% in Belgium, 89.6% in Italy; **Figure 1B**). More than half of the participants declared they would test twice after a first negative result if symptoms still persisted (50.9% in France, 56.7% in Belgium, 68.2% in Italy), whereas few would repeat the test at least a third time (5.8% in France, 12.2% in Belgium, 15.7% in Italy). If access to the test was at a given cost (<20 Euros), then only 42.2% of individuals in France and 47.2% in Belgium would get tested in the presence of symptoms, compared to 72.5% in Italy (**Figure 1C**). Lower values were found if individuals were asymptomatic.

**Figure 1.**
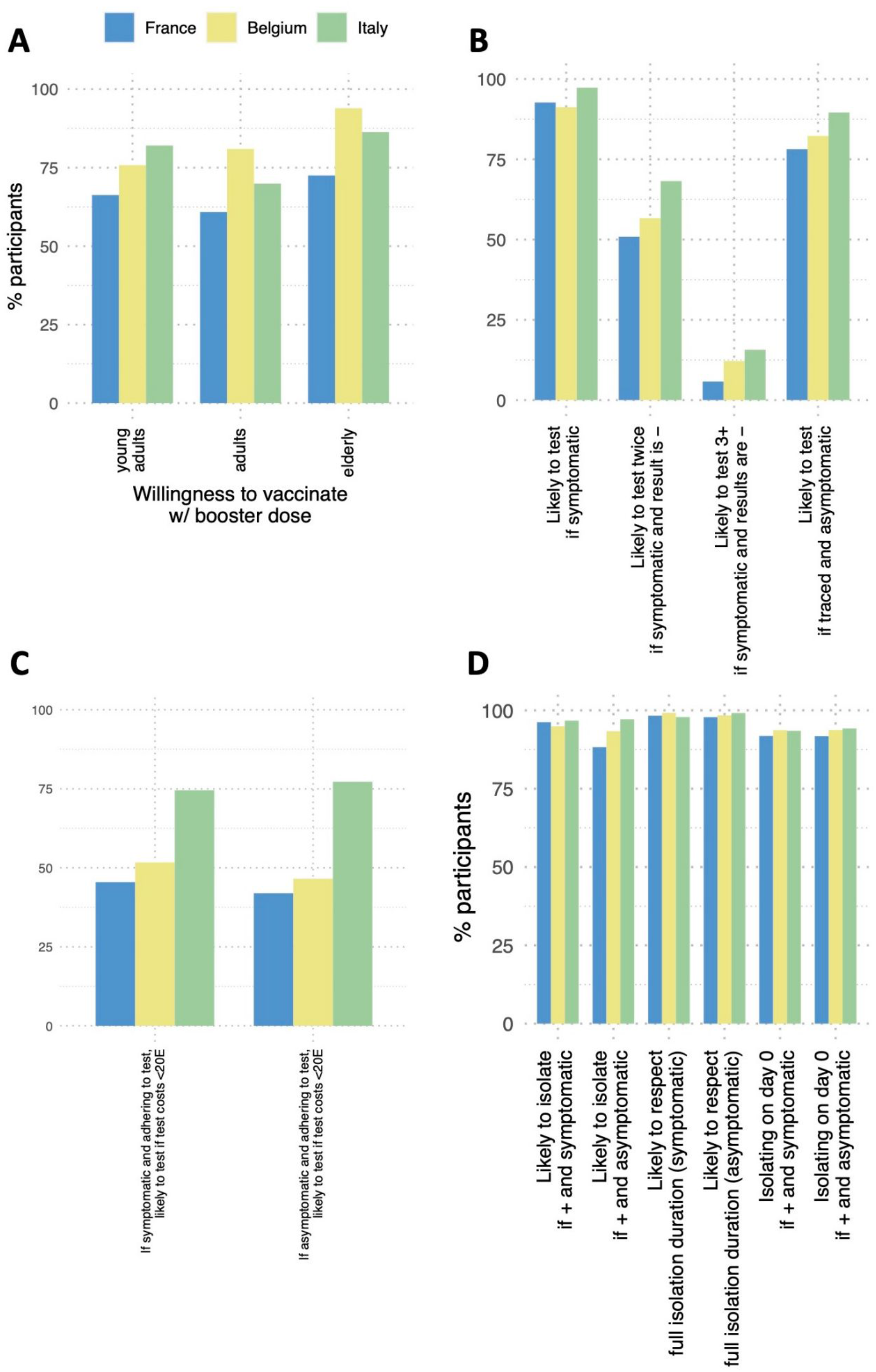
Survey responses on propensity to vaccinate, testing and isolate in the 2022/2023 winter. A: Willingness to vaccinate with a booster dose in the fall 2022 campaign, if recommended, by age class and by country. Age classes correspond to (young adults, adults, elderly, respectively): [20, 39], [40, 59], 60+ for the French and Italian data; [25, 44], [45, 64], 65+ for the Belgian data. B: Adherence to testing during the 2022/2023 winter wave, if recommended, by country. C: Adherence to testing during the 2022/2023 winter wave, if recommended, by country, if tests cost less than 20 euros. D: Adherence to isolation during the 2022/2023 winter wave, if recommended, by country. Responses on isolation duration and day of entry into isolation are conditional to a positive response about isolation.

Participants from all countries reported a high propensity to comply with recommendations for rapid isolation if symptomatic (96.2% would isolate in France, and among them 91.8% would enter isolation on the day of symptoms onset, corresponding to 88% of French respondents; similar values were obtained for Belgium and Italy; **Figure 1D**). Lower values were reported in France for asymptomatic infections compared to the other two countries (88.3% would isolate if asymptomatic in France vs. 93.4% in Belgium and 97.2% in Italy). If isolated, almost full compliance to recommendations on the isolation duration was declared by participants in the three countries (98.3% in France, 99.2% in Belgium, 97.9% in Italy), with no difference between symptomatic and asymptomatic infection.

### Impact of expected booster uptake, adherence to testing and isolation protocols

The fall 2022 booster campaign with declared adherence was estimated to allow the population of each country to restore, during the winter 2022/2023, levels of vaccine-conferred immunity higher than those achieved during the summer of 2022, when the last booster campaign occurred (**Figure 2**). Most importantly, as immunity decreases over time, these levels would be substantially higher than those estimated for the start of the winter 2022/2023 in the absence of the booster campaign. According to senior respondents’ willingness to get vaccinated with a booster, the mean vaccine-conferred protection against symptomatic infection inferred for the overall population of a given country would increase from 3.3% (standard deviation (std) 0.3%) before the booster to 13% (std 4%) after the booster in France (i.e. +294%), from 2.4% (std 0.2%) to 13% (std 5%) in Belgium (+442%), and from 3.8% (std 0.3%) to 16% (std 6%) in Italy (+321%).

**Figure 2.**
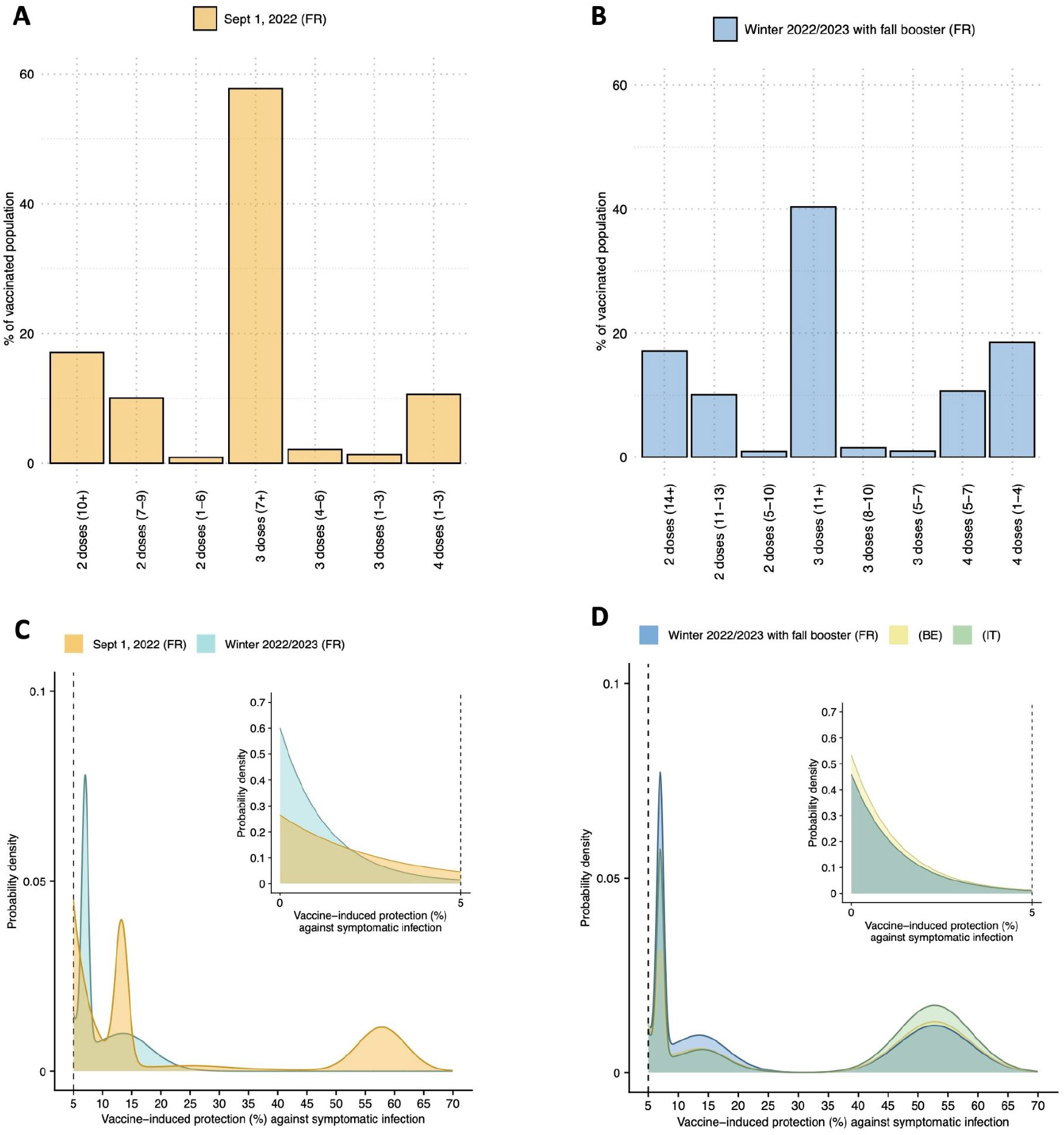
Estimated vaccine-induced immunity in the population. A: Percentage of vaccinated individuals (2+ doses) in the French population in each waning category (time periods expressed in months), as of Sept. 1, 2022. B: Expected percentage of vaccinated individuals (2+ doses) in the French population in each waning category, with a fall booster informed by French survey respondents. C: Density of probability of population-level vaccine-induced protection against symptomatic infection inferred for September 1, 2022, and for the start of the 2022/2023 winter in the absence of the fall 2022 booster campaign. Results refer to France. D: Density of probability of population-level vaccine-induced protection against symptomatic infection inferred for the start of the 2022/2023 winter in the presence of the fall 2022 booster campaign. Adherence to the campaign is informed by the InfluenzaNet survey responses for France, Belgium and Italy.

As the vaccine-induced immunity profiles of the populations, the testing protocols and the prospective adherence to such protocols differed among the three countries under study, we first focused on the French population, and evaluated the expected impact of the three national protocols as if they were applied in France. We used a branching process mathematical model for SARS-CoV-2 diffusion parameterized to Omicron subvariants circulating in France with an effective reproductive number R=1.6 in the absence of interventions (see Methods). Assuming a 75% reduction in transmissibility during the isolation period, we predicted that testing and isolation would reduce R by 18.4% (95% bootstrap confidence interval (CI) 18.1-18.8%) with the French protocol. If, instead, French authorities were to adopt the Belgian or Italian protocol, that would reduce R by 17.2% (16.8-17.4%) and 24.4% (24.0-24.9%), respectively (**Figure 3A**). This corresponds to the Belgian protocols being 2% less effective than the French one, and the Italian protocol being 7% more effective (**Figure 3B**). Given R=1.6, none of the protocols alone would therefore be enough to fully control Omicron spread by reducing the effective reproductive number below 1. These results were based on the expected adherence to testing and isolation declared by French participants in the survey (**Figure 1**) given current access to testing: those vaccinated may get tested at any pharmacy and clinic free of charge, and without prescription (**Table 1**). Negligible differences were estimated when comparing currently active protocols with those in place last winter (2021/2022 winter, **Figure 3**). If tests were to cost up to 20 Euros, the decline in adherence to testing declared by survey participants would be responsible for approximately 60% lower predicted effectiveness of the three testing strategies compared to free access (**Figure 3A**).

**Figure 3.**
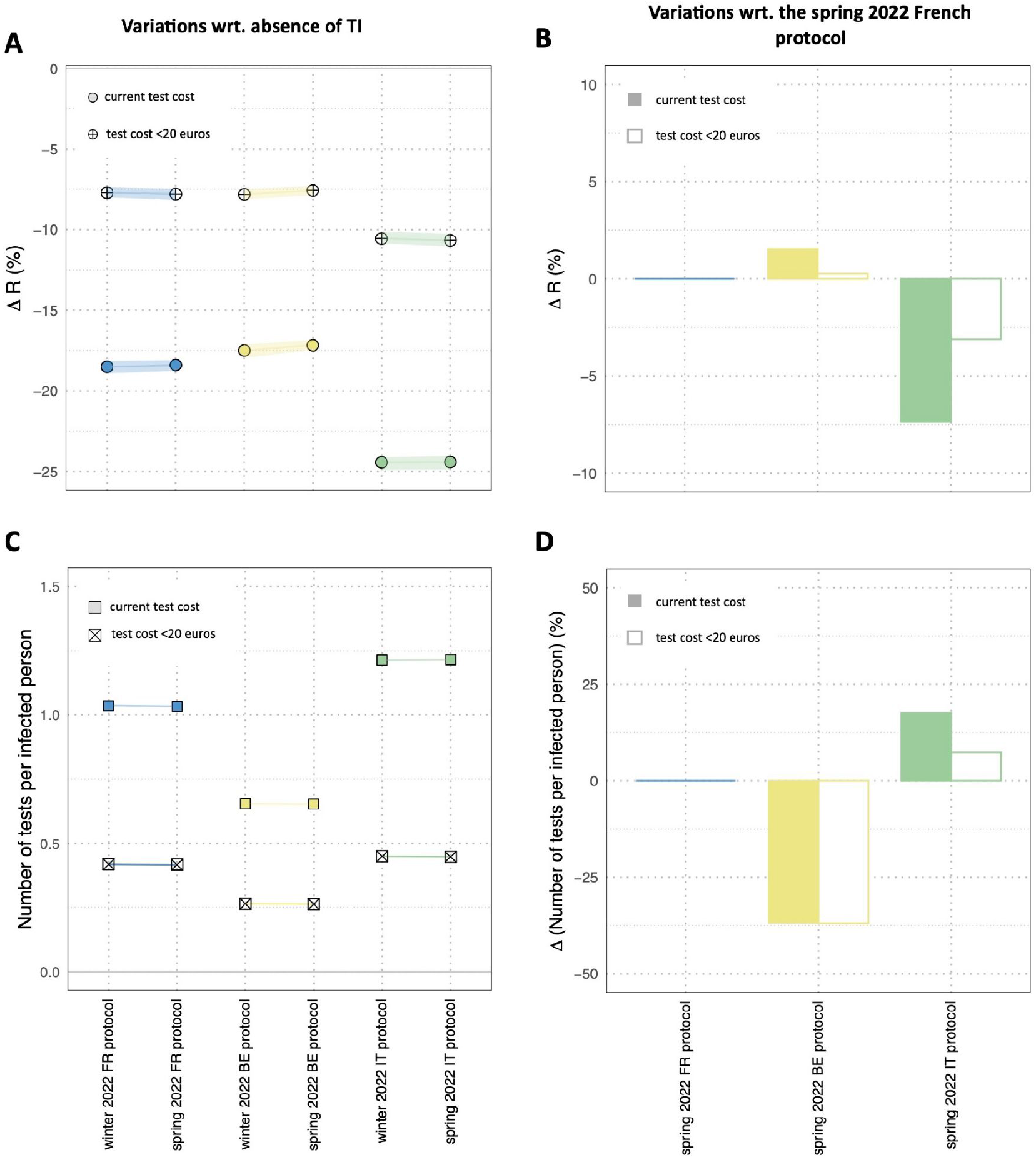
Expected performance of the French, Belgian, and Italian protocols in the 2022/2023 winter. A: Relative variation of the effective reproduction number compared to the no intervention case (absence of TI, testing and isolation) for each protocol. Medians and 95% bootstrap confidence interval. B: Relative variation of the effective reproduction number compared to the spring 2022 French protocol (currently applied). Medians and 95% bootstrap confidence interval. C: Mean number of diagnostic tests per infected case. D: Mean relative variation of the number of diagnostic tests per infected case compared to the spring 2022 French protocol (currently applied). All results refer to the three protocols as if applied in France.

The three protocols implied different testing resources, with on average 1 diagnostic test per infected person using the French protocol, compared to 0.7 and 1.2 per infected person using the Belgian and Italian protocols, respectively (**Figure 3D**). These testing needs would be reduced approximately by 60% if a moderate cost was introduced.

Small variations in the effectiveness and costs of protocols were observed when each testing and isolation protocol was evaluated as applied in its corresponding country, i.e. considering the adherence to interventions declared by the country’s participants, under the same scenario of R=1.6 (**Figure S8** of the Supplementary Information). The higher willingness of Italian survey participants to get tested even if they have to pay out-of-pocket (74.6% vs. 51.7% in Belgium and 45.5% in France if symptomatic, **Figure 1C**) would however increase the predicted effectiveness of the Italian protocol in this context compared to the other countries (19% reduction of R considering the Italian adherence to pay for testing, **Figure S8**, vs. 11% considering the French adherence, **Figure 3**).

A higher isolation effectiveness (90%) would improve the control of the winter wave leading to a 21-29% reduction of community transmission across the three countries (**Figure S10**), however still leaving the effective reproductive number larger than 1 under the R=1.6 scenario considered here. A less effective isolation of infectious individuals (60% reduction of the viral transmissibility) would lower control, yielding a mitigation effort in the range of 15-22% reduction of R (**Figure S9**).

Despite different rules on exit from isolation, the French and Belgian protocols were predicted to impose a similar average length of isolation (about 6 days, after accounting for delays in entry and possible anticipated exits, as declared by survey participants), while releasing similarly high percentages of individuals while still infectious (approximately 60%; **Figure 4**). This percentage would be much smaller (8%) with the Italian protocol because of the higher probability that individuals would isolate for longer (average isolation length of about 11 days). The predicted fraction of infected individuals under isolation exiting after 7 days was indeed 13%, 0%, 68% in the French, Belgian, and Italian protocols, respectively. As a consequence, the predicted share of post-isolation transmission was predicted to be around 19% in the French and Belgian protocols, while it was significantly lower in the Italian protocol (around 8%; **Figure S5**).

**Figure 4.**
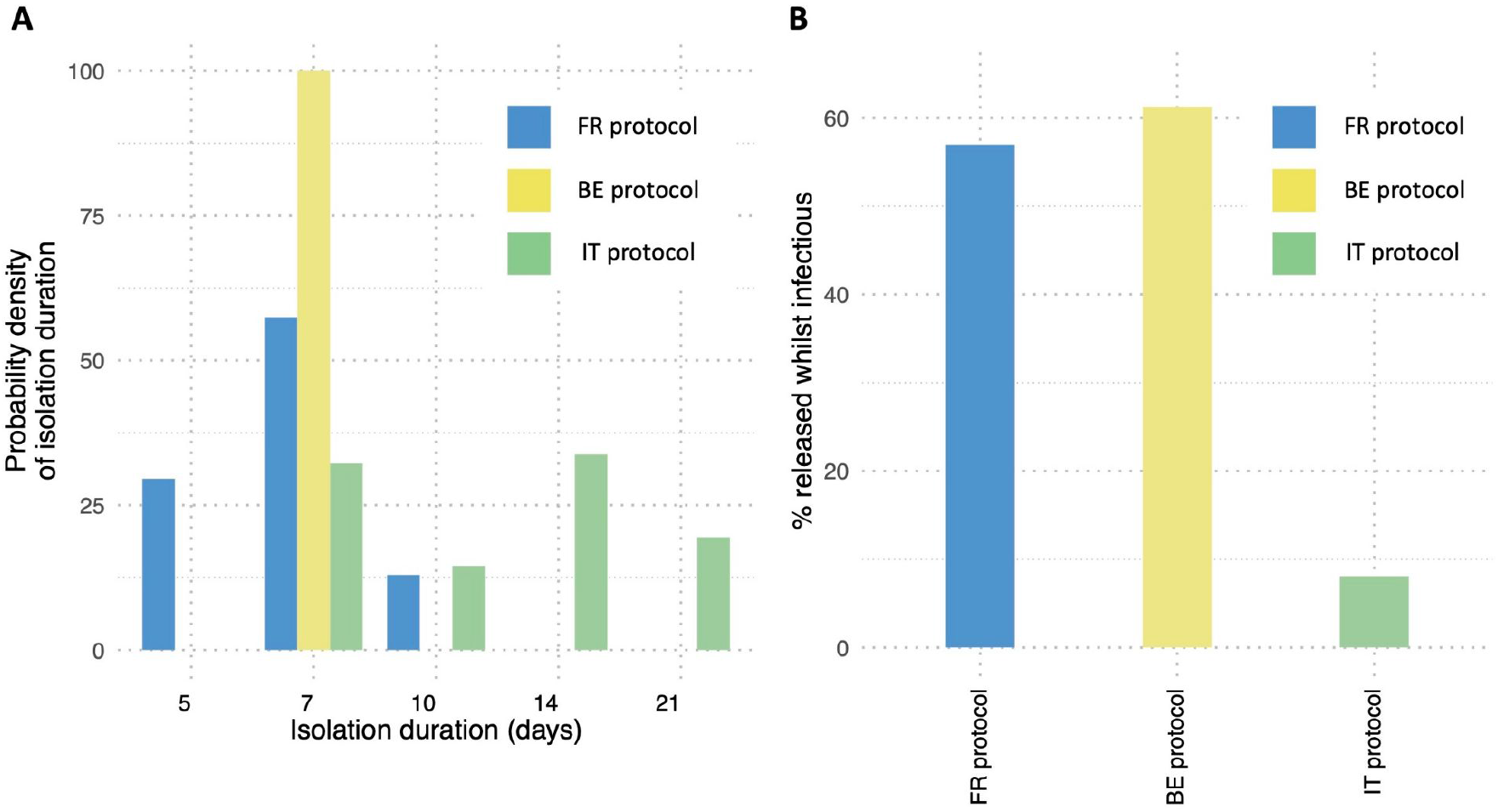
Impact of the duration of isolation. A: Probability density of isolation duration in the French, Belgian, and Italian protocols. B: Percentage of individuals who are released from isolation while still infectious in the French, Belgian, and Italian protocols. All results refer to the three protocols as if applied in France.

## DISCUSSION

While contact tracing has been largely abandoned, several European countries still maintain testing and isolation protocols to mitigate COVID-19 in the next phase of the pandemic. Adapting these strategies to this shifting context means accounting for the expected compliance of a population that is increasingly more exhausted by the health crisis^11,12^. Using responses from 4,594 individuals of a multi-country survey in Europe on their prospective adherence to testing and isolation, our study predicted that current protocols applied in France and Belgium would contribute with a 17-18% reduction of community transmission in the winter wave caused by the Omicron subvariants. Simpler guidelines on isolation duration would decrease the needed testing resources without affecting the performance of these protocols. The Italian protocol would be 7% more effective than the others, but at a higher social cost in terms of days lost in isolation. The protocols alone would however not be enough to bring R below 1 even in the case of the high adherence declared by survey participants. Lower adherence to testing and isolation, due to a lower perceived need of such public health measures or because of the introduction of a cost barrier, would considerably decrease the mitigating impact of these measures. In any scenario, achieving high coverage of the fall booster vaccine dose remains imperative to obtain effective control of the winter wave.

Among the three national protocols studied here, the Italian protocol yielded the largest reduction of the effective reproductive number (24%) compared to the French and Belgian protocols (17-18%), for a moderate effectiveness of isolation (75%). This is due to a longer mandated isolation period, and in particular to isolation being constrained to fixed weekly durations in case of positive exit test. For example, according to the Italian protocol implemented in April 2022^13^, vaccinated individuals with a positive exit test at day 7 were required to continue isolation until day 14, and were allowed to exit only with a negative exit test; otherwise, they had to remain in isolation for an additional week. Only the modifications to the protocol further applied in the fall 2022^13^ relaxed the constraint of the weekly wait, allowing individuals to exit with a negative test after the initial mandatory isolation period. For a 7% loss in mitigating effects, the French and Belgian protocols would ensure shorter isolations - a desirable outcome given the toll of pandemic fatigue building over almost three years^11,12^. Moreover, adopting simpler guidelines on isolation as in the Belgian protocol (with a fixed isolation duration for all individuals) would yield similar durations of isolation compared to the French protocol, but 30% fewer tests required per infected person. This is due to the absence of the exit test that is instead required in France^14^ to shorten isolation (on day 5 if vaccinated, on day 7 if unvaccinated). Simplifying the rules on isolation has the potential to (i) largely reduce the huge costs of the surveillance system^5^ or the costs for households purchasing self-tests; and (ii) encourage higher compliance among pandemic-weary individuals through clear communication of a simplified message^15,16^.

The changes of protocols that occurred in the three countries from winter 2021/2022 to spring 2022 mainly simplified and relaxed the rules on contact tracing, without affecting the requirements for isolation. This change was motivated by the high economic costs of the contact tracing system^5^ and the limited adherence observed in the population^17^ - e.g. not declaring contacts or sharing details of contacts. Our findings predicted that this protocol change has a negligible impact on community transmission.

Earlier modeling works^18,19^ predicted a larger potential of testing and isolation to reduce onward transmission compared to our estimates, e.g. larger than 30%. However, these studies focused on an early phase of the pandemic and assumed perfect adherence to isolation and 100% isolation effectiveness. In contrast, in the current phase, greater fatigue and an evolving perception of the risk will likely affect these two aspects. Our main findings were obtained considering the responses on prospective attitudes towards testing and isolation provided by the survey participants in the three countries. Survey data were adjusted to account for the socio-demographic differences observed between the Influenzanet digital cohorts and the corresponding national populations. Previous work showed that adjusted trends of estimated influenza-like-illness incidence from Influenzanet reports compare well with those of national sentinel systems^8,20^, proving the accuracy of the Influenzanet participatory system despite its lack of representativeness. However, we cannot exclude that the cohort under study suffers from self-selection biases towards increased interest in health topics that could possibly also boost willingness to comply with preventive behaviors. Data from 37 nationally representative surveys in the UK between March 2020 and January 2021 reported that only 20% of respondents would seek a diagnostic test if they had symptoms, and that duration-adjusted adherence was around 50%^21^. These behaviors may however be context-specific and induced by the financial and practical support in place^22^ (or lack thereof), so that they can hardly be generalizable elsewhere. In addition, support and access varied over time. For example, in the period following the first wave in France, seeking a general practitioner to obtain a mandatory prescription to testing was a major barrier to testing and isolation undermining the effectiveness of the surveillance and control system^9^, and was later lifted. Similarly, in our multicountry survey we found that adherence to testing would be reduced by more than half in French and Belgian respondents if tests were to cost up to 20 Euros, even in the presence of COVID-like symptoms. This would halve the effectiveness of testing and isolation in reducing community transmission, showing that such policy change may substantially affect the capacity to control the winter wave. In contrast, a larger fraction of Italian survey participants would agree to test despite the cost barrier. This may possibly be due to habits developed in the Italian context with individuals buying tests at the pharmacies to avoid lengthier and less directly accessible procedures to access free tests through local health authorities. Providing adequate and comprehensive support, including e.g. paid sick leave to encourage and enable self-isolation when sick, remains an important aspect for the management of COVID-19 even beyond the pandemic phase. This would allow the reduction of health disparities^23^ and of the overall costs of absenteeism^24^ from work due to the spread of winter infections.

While prospective compliance to testing and isolation was rather homogeneous across countries, with the few exceptions discussed above, the willingness to get the booster dose recommended to at-risk individuals in the fall 2022 was markedly different. Among senior respondents, we found a higher propensity to vaccinate with the booster dose in Belgium, followed by Italy, and then France, with an uptake overall larger than 72%. The actual coverage reported by early December 2022 for the fall booster campaign that started in October-November 2022, varies considerably across countries: 11% in France^25^, 52% in Belgium^26^, 17% in Italy^27^ in seniors. Fall booster coverage in adults was negligible (e.g., <2% in France^25^ as of December 7, 2022). While the authorities and the scientific community continue to support recommendations to get vaccinated with a booster dose in prevention for the winter wave, it is possible that the changing threat with respect to the previous two winters is deterring individuals. A change in the message delivery, from central authorities to local health care providers, GPs, and pharmacists may help in boosting vaccination choices. Direct recommendations from healthcare workers have proven to be a strong determinant of vaccination uptake against influenza^28^, improving the quality of information provided and increasing patients’ trust. Compared to the scenario considered in the main analysis, the low vaccination coverages reported so far in France and in Italy would correspond to a higher effective reproductive number, compromising the ability of testing and isolation strategies to slow down the case surge in the winter wave, as shown in our sensitivity analysis. Waiting for possible increases of vaccination uptake over the winter, preventive measures such as masking^29^ in closed settings become again essential to reduce onward transmission.

This study has a set of limitations. First, the model is not age-stratified. This is due to the branching process approximation, typically used to study the effects of testing, tracing and isolation in detail^30–34^. However, we do consider the age-specific response to booster uptake in the multi-country survey to infer the population-level vaccine-induced immunity for the 2022/2023 winter. Second, our approach does not consider the healthcare impact in terms, for example, of hospital admissions and saturation. Similarly to the first limitation, this is related to the modeling framework and the absence of age stratification that characterizes the severity of the infection. Though our study does not characterize the epidemiological conditions putting pressure on the healthcare system, its findings can be used to anticipate the expected reduction of the effective reproductive number to slow down the viral circulation, with consequent effects on the hospital admissions. Finally, we fixed the effectiveness of isolation to 75%, corresponding to a 75% reduction of transmissibility while the individual is isolated. Previous works^18,19^ generally assumed higher values, including full compliance, but they also referred to the acute phase of the pandemic when more severe variants circulated in a less immunized population, and non-pharmaceutical interventions were the key response to the pandemic^35^. In that context, high compliance to isolation to reduce contacts within households was expected. In a post-crisis phase, where individuals received several vaccine doses and were possibly exposed to previously circulating variants, it is plausible to assume a lower effectiveness of isolation. In the sensitivity analysis we showed that higher values of isolation effectiveness would increase the resulting mitigating impact on COVID-19 circulation, as expected, while preserving the overall conclusions.

As European countries enter their third winter since SARS-CoV-2 emergence with a generalized feeling that the pandemic is over^3^, it is important to evaluate in this shifting context the tradeoff between applicable mitigation measures and the expected compliance to them. Our findings show that simpler guidelines for testing and isolation protocols, similar to the Belgian protocol, would be as efficient as more elaborate guidelines (e.g. the French protocol) while reducing the cost of testing resources needed. Longer isolation periods found in the Italian protocols may not be sustainable in this phase of the pandemic. Booster uptake remains a fundamental element of the response to manage COVID-19 through spring 2023.

## METHODS

### Influenzanet multicountry survey and analysis

We used the existing European platform Influenzanet for online participatory surveillance of influenza-like-illness in the general population^8^, adapted to COVID-19 during the pandemic crisis^9,10,36^. Three countries participated in the study: France, Belgium and Italy. Each country ran its own website, to which participants registered on a voluntary basis, completing an intake survey covering demographical, socio-economic and health-related factors. An email was sent to participants with a personal link to the survey on prospective vaccination, testing and isolation, on June 25, 2022 in France, July 7 in Italy, July 14 in Belgium. Data collection was closed on July 1, July 18, July 25 in the three countries, respectively. In case of multiple submissions by a single participant, only answers submitted last were considered in the study. Participants who indicated their gender, age, education level, and vaccination status were included in the study, provided their age met the age/gender/education stratification available for each country (see below). “Undefined” gender type in Belgium was excluded from the analysis. The survey text is provided in the Supplementary Information.

The Influenzanet sample differed from the general population (**Table S1** of the Supplementary Information). We therefore adjusted the French, Italian, and Belgian survey data on age, gender, and level of education, as in previous work^10,37^, using data from the French National Institute of Statistics and Economic Studies (Institut National de la Statistique et des Études Économiques, INSEE^38^), the Belgian National Institute of Statistics and Census Data (STATBEL^39^), and the Italian National Institute of Statistics (Istituto nazionale di statistica, ISTAT^40^). Based on available data from these sources, we considered the following age classes: [20, 39], [40, 59], 60+ for France and Italy; [25, 44], [45, 64], 65+ for Belgium. For the level of education, we considered the categories: <=A-levels and >A-levels (the full correspondence with the educational levels in each country is provided in the SI, **Table S3**). Survey participants who did not belong to any of these classes were excluded from the study.

### Ethics statement

GrippeNet.fr/COVIDnet.fr was reviewed and approved by the French Advisory Committee for research on information treatment in the health sector (CCTIRS), and by the French National Commission on Informatics and Liberty (CNIL) - the authorities ruling on all matters related to ethics, data, and privacy in France. Infectieradar.be was reviewed and approved by the Committee for Medical Ethics of Antwerp University (EC UZA-UA). Influweb.it was reviewed and approved by the IRB of the ISI Foundation, its use takes place in compliance with the rules contained in the GDPR. Informed consent was obtained online from all participants of the three platforms at enrollment according to regulations, enabling the collection, storage, and treatment of data, and their publication in anonymized, processed, and aggregated forms for scientific purposes. All three websites have a “Privacy Statement” section in which the users who decide to enroll in the study can find all the information on who is responsible for the data acquisition and processing in each country.

### Model structure, testing and isolation

We adapted a branching process model previously introduced by Hellewell et al.^31^. The model distinguished between symptomatic and asymptomatic cases, and considered their vaccination status. The number of secondary cases generated by each infected individual was drawn from a negative binomial distribution to account for the large individual variation observed in SARS-CoV-2 transmission^41–45^. The mean of the distribution was set to the chosen value of the reproductive number, in the absence of testing and isolation (see the following subsection). We parameterized the model with the distributions of the generation time and of the incubation period estimated for the Omicron (B.1.1.529) subvariant of SARS-CoV-2^46^. Each potential new case was assigned a time of infection drawn from the distribution of generation time, the possibility to develop symptoms^47,48^, and a time of symptoms onset drawn from the incubation period distribution. The number of secondary cases accounted for the reduced infectiousness of asymptomatic individuals^49^ and the vaccine effectiveness against transmission^50^. We considered the vaccine effectiveness against Omicron symptomatic infection^51^ in reducing the probability to develop symptomatic forms of the disease, and considered that the vaccine effectiveness against infection was 10% lower. The estimates for the vaccine effectiveness accounted for the number of doses and the waning in time since the last administration (see next subsection). The vaccine-induced immunity profile of the population in the model was inferred from the reported vaccine coverage and the timing of the vaccination campaigns in each of the three countries considered (see next subsection). Natural immunity from prior SARS-CoV-2 infections and hybrid immunity were considered by setting the value of the effective reproductive number of the winter wave.

Infected individuals in the model could enter isolation because they tested positive (see the last subsection of the Methods). We considered adherence to testing if symptomatic, in the presence of recommendations to do so, parameterized with the responses of the Influenzanet survey participants. Those not testing or testing negative did not enter isolation. Individuals testing negative in the presence of symptoms in the model were allowed to repeat the test one or more times, as we considered a certain level of awareness regarding test sensitivity after symptoms onset^52^. This awareness was informed by survey responses. Individuals testing positive (at any of these tests after symptoms onset) could isolate themselves, according to the adherence to isolation recorded in the survey in each country. We allowed for an onset-to-isolation delay drawn from an exponential distribution with average value taken from survey responses. We considered a 75% reduction of transmission in isolation, close to the estimates of Ref.^53^ and considering that isolation may be less effective during the third COVID-19 winter because of pandemic fatigue^11,12^ and a lower perceived risk of infection and severe outcome due to widespread vaccination^54^. We also explored 60% and 90% isolation effectiveness for sensitivity. Some protocols required a negative exit test to terminate isolation, and in the model we simply assumed full adherence to the exit test. However, individuals could exit isolation before its prescribed duration and we informed this probability from survey responses in each country, considering an anticipation with respect to the due date drawn from an exponential distribution with average value being a fixed fraction (3/4) of the recommended isolation duration. We considered lateral flow tests and their temporal diagnostic sensitivity to reproduce the observed rate of LFT test positivity over time after an Omicron infection^52,55^ (**Figure S3**).

We assumed that in the 2022/2023 winter contact tracing will not be adopted anymore by authorities, as it is largely costly in terms of resources and carries a heavy social impact due to the imposed quarantine^5,6^. However, we considered a residual informal contact tracing self-performed by individuals who would alert their known contacts (relatives, friends, co-workers) that they tested positive to COVID-19. We assumed that this residual contact tracing amounts to tracing 5% of the occurred infections.

A visualization of the branching process model is provided in **Figure S1** of the Supplementary Information. Model parameters and sources are provided in **Table S3**.

### 2022/2023 winter COVID-19 scenarios

We considered a winter wave due to Omicron subvariants occurring in the three countries characterized by a reproductive number *R*=1.6 in the absence of testing and isolation^56^. Additional values of *R* were explored in the sensitivity analysis. Vaccine-induced immunity profile of the population of each country was estimated based on vaccination coverage reported in France^25^, Belgium^57^, and Italy^27^ up to summer 2022, immunity waning, and uptake of the fall 2022 booster campaign for the elderly as declared by Influenzanet survey participants. We considered estimates of vaccine effectiveness against symptomatic infection from the Omicron variant^51^ for the Pfizer-BioNTech BNT162b2 vaccine, which was predominantly used in France, Belgium and Italy in the COVID-19 vaccination campaign. To account for the reduction of vaccine protection over time, we considered the following waning process^51^: 2 vaccine doses at 1-6 months since the second dose, at 7-9 months, and at 10+ months; 3 vaccine doses at 1-3 months since the third dose, 4-6 months, or 7+ months; 4 vaccine doses within 3 months from the fourth dose. From the reported coverage in each country on September 1, 2022 we estimated the probability density of vaccine-induced protection against symptomatic infection for the French, Belgian, and Italian populations at that date (**Figure 2**). Applying 4 months of waning, we estimated this probability density at the start of the 2022/2023 winter. Considering the share of the elderly survey respondents willing to get vaccinated with the fall 2022 booster, we computed the expected vaccine-induced protection in each population given the reported coverage and the prospective uptake of the fall booster dose in each country at the start of the 2022/2023 winter.

### Testing and isolation protocols

We implemented the testing and isolation protocols adopted by France, Belgium, and Italy in the 2021/2022 winter and spring 2022^13,58,59^ (**Table 1**). In all protocols, symptomatic individuals were required to test immediately upon symptom onset and to isolate, if positive. Protocols differed in the duration of the isolation periods and in the exit test strategies. For example, last winter protocols required 5 days with a negative exit test and absence of symptoms in the last 48h or 7 days (with no test) in France, 7 days (with no exit test) in Belgium, 7 days with a negative exit test or 14 days with a negative exit test or 21 days (with no test) in Italy. Other differences referred to distinct isolation mandates depending on vaccination status (in France and in Italy), and to protocols for contact tracing impacting the testing of contacts and their quarantine duration.

In all countries, spring 2022 protocols relaxed certain restrictions foreseen by previous protocols, notably simplifying the contact tracing. As of December 2022, France and Belgium still adopt the spring 2022 protocols (in Belgium, with some regional variations), whereas Italy revised it in early September 2022 (see **Table 1**), in particular to reduce the length of isolation up to 5 days with a negative exit test, or 14 days with no test in case of persistent positivity, for both vaccinated and unvaccinated individuals^13^. As this last protocol is rather similar to the French spring protocol, we considered in the study the 3 protocols adopted during spring 2022 and commented on the implications of this change in the Discussion section.

## Data Availability

Survey individual data cannot be shared owing to restrictions imposed by the national data protection authorities. Stratified data on survey responses will be made available in a persistent repository upon publication.

## ACKNOWLEDGEMENTS

We thank all participants in the surveys. We thank Daniel Levy-Bruhl, Isabelle Parent, and Isabelle Bonmarin for useful comments and discussions on this study. This study was partly supported by: Agence Nationale de la Recherche projects COSCREEN (ANR-21-CO16-0005) and DATAREDUX (ANR-19-CE46-0008-03) to VC; ANRS–Maladies Infectieuses Émergentes project EMERGEN (ANRS0151) to VC; EU Horizon 2020 grants MOOD (H2020-874850) and RECOVER (H2020-101003589) to VC, EpiPose (101003688) to DP, LH, NH; EU Horizon Europe VERDI (101045989) to VC, DP; the Chaire Blaise Pascal Program of the Île-de-France region to JSW. Views and opinions expressed are those of the authors and do not necessarily reflect those of the funding agencies and authorities, who cannot be held responsible for them.

## AUTHORS’ CONTRIBUTION

VC conceived and designed the study. GdM and VC designed the initial survey draft. EV, CC, MD, CKK, HN, DP, LH, NH contributed to revising and finalizing the survey. MD, CKK, CT, LH, DP implemented the survey in the national Influenzanet platforms. GdM developed the code and analyzed the data. All authors interpreted the results. GdM and VC wrote the initial manuscript draft. All authors edited and approved the final version of the Article.

## REFERENCES

1. Copenhagen: WHO Regional Office for Europe. Strategy considerations for severe acute respiratory syndrome coronavirus 2 (SARS-CoV-2) and other respiratory viruses in the WHO European Region during autumn and winter 2022/23: protecting the vulnerable with agility, efficiency, and trust. https://www.who.int/europe/publications/i/item/WHO-EURO-2022-5851-45616-65461 (2022).

2. COVID-19: the next phase and beyond. The Lancet 399, 1753 (2022).

3. Carlos del Rio, MD & Preeti N. Malani. COVID-19 in 2022—The Beginning of the End or the End of the Beginning? | Infectious Diseases | JAMA | JAMA Network (2022) doi:10.1001/jama.2022.9655.

4. Looi, M.-K. Is covid-19 settling into a pattern? BMJ 378, o2183 (2022).

5. Cour des comptes. Tracer les contacts des personnes contaminées par la covid 19. https://www.ccomptes.fr/fr/publications/tracer-les-contacts-des-personnes-contaminees-par-la-covid-19.

6. Bonati, M., Campi, R. & Segre, G. Psychological impact of the quarantine during the COVID-19 pandemic on the general European adult population: a systematic review of the evidence. Epidemiol. Psychiatr. Sci. 31, e27 (2022).

7. European Centre for Disease Prevention and Control. Preliminary public health considerations for COVID-19 vaccination strategies in the second half of 2022. https://www.ecdc.europa.eu/en/publications-data/preliminary-public-health-considerations-covid-19-vaccination-strategies-second (2022).

8. Guerrisi, C. et al. Participatory Syndromic Surveillance of Influenza in Europe. J. Infect. Dis. 214, S386–S392 (2016).

9. Pullano, G. et al. Underdetection of cases of COVID-19 in France threatens epidemic control. Nature 590, 134–139 (2021).

10. McColl, K. et al. Are People Optimistically Biased about the Risk of COVID-19 Infection? Lessons from the First Wave of the Pandemic in Europe. Int. J. Environ. Res. Public. Health 19, 436 (2022).

11. The New York Times. Opinion | The New Phase of the Pandemic Is Covid Exhaustion. https://www.nytimes.com/2022/03/09/opinion/covid-exhaustion-the-argument.html.

12. Petherick, A. et al. A worldwide assessment of changes in adherence to COVID-19 protective behaviours and hypothesized pandemic fatigue. Nat. Hum. Behav. 5, 1145–1160 (2021).

13. Ministero della Salute. Test diagnostici, contact tracing, isolamento e autosorveglianza. https://www.salute.gov.it/portale/nuovocoronavirus/dettaglioFaqNuovoCoronavirus.jsp?lingua=italiano&id=244#7.

14. ameli.fr. Symptômes, gestes barrières, cas contact et isolement. https://www.ameli.fr/assure/covid-19/symptomes-gestes-barrieres-cas-contact-et-isolement.

15. Hung, L. & Lin, M. Clear, consistent and credible messages are needed for promoting compliance with COVID-19 public health measures. Evid. Based Nurs. 25, 22–22 (2022).

16. Rubin, R. How Immune-Evasive Omicron Offspring and a Lack of Mitigation Measures Could Shape a COVID-19 Winter Wave. JAMA 328, 2092–2095 (2022).

17. Le Monde. « Ne donne pas mon nom » : le traçage des cas contacts de malades du Covid-19 patine. https://www.lemonde.fr/les-decodeurs/article/2021/08/22/ne-donne-pas-mon-nom-le-tracage-des-cas-contacts-de-malades-du-covid-19-patine_6092034_4355770.html.

18. Grassly, N. C. et al. Comparison of molecular testing strategies for COVID-19 control: a mathematical modelling study. Lancet Infect. Dis. 20, 1381–1389 (2020).

19. Wells, C. R. et al. Optimal COVID-19 quarantine and testing strategies. Nat. Commun. 12, 356 (2021).

20. Guerrisi, C. et al. The potential value of crowdsourced surveillance systems in supplementing sentinel influenza networks: the case of France. Eurosurveillance 23, 1700337 (2018).

21. Louise E Smith et al. Adherence to the test, trace, and isolate system in the UK: results from 37 nationally representative surveys | The BMJ. BMJ 372, (2021).

22. Thorneloe, R. J., Clarke, E. N. & Arden, M. A. Adherence to behaviours associated with the test, trace, and isolate system: an analysis using the theoretical domains framework. BMC Public Health 22, 567 (2022).

23. Zipfel, C. M., Colizza, V. & Bansal, S. Health inequities in influenza transmission and surveillance. PLOS Comput. Biol. 17, e1008642 (2021).

24. Asfaw, A., Rosa, R. & Pana-Cryan, R. Potential Economic Benefits of Paid Sick Leave in Reducing Absenteeism Related to the Spread of Influenza-Like Illness. J. Occup. Environ. Med. 59, 822–829 (2017).

25. Données relatives aux personnes vaccinées contre la Covid-19 (VAC-SI) - data.gouv.fr. https://www.data.gouv.fr/fr/datasets/donnees-relatives-aux-personnes-vaccinees-contre-la-covid-19-1/.

26. Sciensano et al. COVID-19 - BULLETIN EPIDEMIOLOGIQUE DU 20 DECEMBRE 2022. https://covid-19.sciensano.be/sites/default/files/Covid19/Derni%C3%A8re%20mise%20%C3%A0%20jour%20de%20la%20situation%20%C3%A9pid%C3%A9miologique.pdf.

27. Governo Italiano - Report Vaccini Anti Covid-19. https://www.governo.it/it/cscovid19/report-vaccini/.

28. Bartolo, S. et al. Determinants of influenza vaccination uptake in pregnancy: a large single-Centre cohort study. BMC Pregnancy Childbirth 19, 510 (2019).

29. Brooks, J. T. & Butler, J. C. Effectiveness of Mask Wearing to Control Community Spread of SARS-CoV-2. JAMA 325, 998–999 (2021).

30. Davis, E. L. et al. Contact tracing is an imperfect tool for controlling COVID-19 transmission and relies on population adherence. Nat. Commun. 12, 5412 (2021).

31. Hellewell, J. et al. Feasibility of controlling COVID-19 outbreaks by isolation of cases and contacts. Lancet Glob. Health 8, e488–e496 (2020).

32. Fyles, M. et al. Using a household-structured branching process to analyse contact tracing in the SARS-CoV-2 pandemic. Philos. Trans. R. Soc. B Biol. Sci. 376, 20200267 (2021).

33. Laha, A. & Majumdar, S. A multi-type branching process model for epidemics with application to COVID-19. Stoch. Environ. Res. Risk Assess. (2022) doi:10.1007/s00477-022-02298-9.

34. Zhang, L. et al. A Heterogeneous Branching Process with Immigration Modeling for COVID-19 Spreading in Local Communities in China. Complexity 2021, e6686547 (2021).

35. Ge, Y. et al. Untangling the changing impact of non-pharmaceutical interventions and vaccination on European COVID-19 trajectories. Nat. Commun. 13, 3106 (2022).

36. McDonald, S. A. et al. Testing behaviour and positivity for SARS-CoV-2 infection: insights from web-based participatory surveillance in the Netherlands. BMJ Open 11, e056077 (2021).

37. McDonald, S. A. et al. Risk factors associated with the incidence of self-reported COVID-19-like illness: data from a web-based syndromic surveillance system in the Netherlands. Epidemiol. Infect. 149, e129 (2021).

38. Insee. FOR2 - Population non scolarisée de 15 ans ou plus par sexe, âge et diplôme le plus élevé en 2019 − France entière −Diplômes - Formation en 2019. https://www.insee.fr/fr/statistiques/6455250?sommaire=6455252&geo=FE-1.

39. Statbel. Datalab Hoogste opleidingsniveau volgens gewest, geslacht, nationaliteit en herkomst. https://statbel.fgov.be/nl/open-data/datalab-hoogste-opleidingsniveau-volgens-gewest-geslacht-nationaliteit-en-herkomst.

40. ISTAT. Grado di istruzione dettagliato della popolazione residente di 6 anni e più. http://dati-censimentopopolazione.istat.it/Index.aspx?DataSetCode=DICA_GRADOISTR1.

41. Adam, D. C. et al. Clustering and superspreading potential of SARS-CoV-2 infections in Hong Kong. Nat. Med. 26, 1714–1719 (2020).

42. Susswein, Z. & Bansal, S. Characterizing superspreading of SARS-CoV-2 : from mechanism to measurement. 2020.12.08.20246082 Preprint at https://doi.org/10.1101/2020.12.08.20246082 (2020).

43. Wang, J. et al. Superspreading and heterogeneity in transmission of SARS, MERS, and COVID-19: A systematic review. Comput. Struct. Biotechnol. J. 19, 5039–5046 (2021).

44. Wang, L. et al. Inference of person-to-person transmission of COVID-19 reveals hidden super-spreading events during the early outbreak phase. Nat. Commun. 11, 5006 (2020).

45. Lloyd-Smith, J. O., Schreiber, S. J., Kopp, P. E. & Getz, W. M. Superspreading and the effect of individual variation on disease emergence. Nature 438, 355–359 (2005).

46. Manica, M. et al. Intrinsic generation time of the SARS-CoV-2 Omicron variant: An observational study of household transmission. Lancet Reg. Health - Eur. 19, 100446 (2022).

47. Lavezzo, E. et al. Suppression of a SARS-CoV-2 outbreak in the Italian municipality of Vo’. Nature 584, 425–429 (2020).

48. Ma, Q. et al. Global Percentage of Asymptomatic SARS-CoV-2 Infections Among the Tested Population and Individuals With Confirmed COVID-19 Diagnosis: A Systematic Review and Meta-analysis. JAMA Netw. Open 4, e2137257 (2021).

49. Li, R. et al. Substantial undocumented infection facilitates the rapid dissemination of novel coronavirus (SARS-CoV-2). Science 368, 489–493 (2020).

50. Gardner, B. J. & Kilpatrick, A. M. Estimates of reduced vaccine effectiveness against hospitalization, infection, transmission and symptomatic disease of a new SARS-CoV-2 variant, Omicron (B.1.1.529), using neutralizing antibody titers. Preprint at https://doi.org/10.1101/2021.12.10.21267594 (2021).

51. UK Health Security Agency. COVID-19 vaccine surveillance report: week 31. 54 https://assets.publishing.service.gov.uk/government/uploads/system/uploads/attachment_data/file/1096327/Vaccine_surveillance_report_week_31_2022.pdf (2022).

52. Landon, E. et al. High Rates of Rapid Antigen Test Positivity After 5 days of Isolation for COVID-19. http://medrxiv.org/lookup/doi/10.1101/2022.02.01.22269931 (2022) doi:10.1101/2022.02.01.22269931.

53. Smith, H. The Rùm Model Technical Annex. 28 https://assets.publishing.service.gov.uk/government/uploads/system/uploads/attachment_data/file/960110/RUM_model_technical_annex_final100221.pdf (2021).

54. Domoslawska-Zylinska, K., Krysińska-Pisarek, M., Czabanowska, K. & Sesa, G. Vaccinated and Unvaccinated Risk Perceptions and Motivations for COVID-19 Preventive Measures Based on EPPM-A Polish Qualitative Pilot Study. Int. J. Environ. Res. Public. Health 19, 13473 (2022).

55. Schrom, J. et al. Comparison of SARS-CoV-2 Reverse Transcriptase Polymerase Chain Reaction and BinaxNOW Rapid Antigen Tests at a Community Site During an Omicron Surge. Ann. Intern. Med. 175, 682–690 (2022).

56. Genomic surveillance of SARS-CoV-2 in Belgium. https://www.uzleuven.be/nl/laboratoriumgeneeskunde/genomic-surveillance-sars-cov-2-belgium.

57. Dashboard Covid Vaccinations Belgium. http://covid-vaccinatie.be/en.

58. Assurance Maladie. Que faire en cas de symptômes évoquant le Covid-19 ? Ameli https://www.ameli.fr/assure/covid-19/symptomes-gestes-barrieres-cas-contact-et-isolement/symptomes-covid-que-faire.

59. info-coronavirus. Contact tracing: slowing down the virus together | Coronavirus COVID-19. https://www.info-coronavirus.be/en/contact-tracing/.

